# Prediction of abnormal left ventricular geometry in patients without cardiovascular disease through machine learning: An ECG-based approach

**DOI:** 10.1101/2020.11.10.20228981

**Authors:** Eleni Angelaki, Maria E. Marketou, Georgios D. Barmparis, Alexandros Patrianakos, Panos E. Vardas, Fragiskos Parthenakis, Giorgos P. Tsironis

## Abstract

Cardiac remodeling is recognized as an important aspect of cardiovascular disease (CVD) progression. Machine learning (ML) techniques were applied on basic clinical parameters and electrocardiographic features for detecting abnormal left ventricular geometry (LVG), even before the onset of left ventricular hypertrophy (LVH), in a population without established CVD. After careful screening, we enrolled 528 subjects with and without essential hypertension, but no other indications of CVD. All patients underwent a full echocardiographic evaluation and were classified into 3 groups; normal geometry (NG), concentric remodeling without LVH (CR), and LVH. Abnormal LVG was identified as increased relative wall thickness (RWT) and/or left ventricular mass index (LVMi). We trained nonlinear predictive ML models, to classify subjects with abnormal LVG and calculated SHAP values to perform feature importance and interaction analysis. Hypertension, age, body mass index over the Sokolow-Lyon voltage, QRS-T angle, and QTc duration were some of the most important features. Our model was able to distinguish NG from all others (CR+LVH), with accuracy 86%, specificity 75%, sensitivity 95%, and area under the receiver operating curve (AUC/ROC) 0.89. We also trained our model to classify NG and CR (NG+CR) against those with established LVH, with accuracy 89%, specificity 97%, sensitivity 50%, and AUC/ROC 0.85. Our ML algorithm effectively detects abnormal LVG even at early stages. Innovative solutions are needed to improve risk stratification of patients without established CVD, especially in primary care settings, and ML may enable this direction.

## INTRODUCTION

Artificial intelligence (AI) and the application of machine learning (ML) algorithms in the management of data are transforming the landscape of different scientific fields, including clinical medicine.^1^ AI techniques have the potential to radically change the way we practice cardiovascular medicine, providing new tools to interpret data and make clinical decisions. While still a new player in cardiology, ML has already made its mark in clinical diagnostics^2,3^ and research^4,5^ and continues to evolve rapidly. AI offers opportunities not only to physicians to make more accurate and prompt diagnoses, but it can also identify hidden opportunities to improve patient management and avoid unnecessary spending.

Cardiac remodeling is considered an important aspect of cardiovascular disease (CVD) progression and is therefore emerging as a significant therapeutic target.^6–9^ More specifically, arterial hypertension is associated with a spectrum of cardiac geometric adaptation matched to systemic hemodynamics and ventricular load, which has important prognostic implications.^6–9^ Abnormal left ventricular geometry (LVG) in hypertensives is frequently associated with diastolic dysfunction, and left ventricular hypertrophy (LVH) is a major adverse prognostic risk factor for cardiovascular events.^10,11,12^ The electrocardiogram (ECG) is one of the most widely used diagnostic tools; it is of paramount importance in the initial evaluation of a patient suspected to have a cardiovascular pathology. However, ECG is not a sensitive method of detecting left ventricular hypertrophy (LVH), and undoubtedly, under the existing knowledge, it cannot detect changes of LVG at early stages, especially before LVH is present. Notably, although the most commonly used ECG criteria demonstrate relatively high specificity, their sensitivity for the detection of LVH is low, approximately 30%^13^ and in some studies it is as low as 6.9%.^14^ For that reason, and given the importance of abnormal LVG, echocardiogram is suggested as an additional diagnostic evaluation to hypertensive patients according to the latest European guidelines for arterial hypertension.^15^

Recently, using digital interpretation of electrocardiogram (ECG) via computational methods and ML applications, we can extract information that is not easily and directly detected by the human eye, especially within a busy clinical setting. On the other hand, echocardiography is a more sensitive approach for the detection of cardiac morphologic changes mediated by arterial hypertension, and more valuable for cardiovascular risk assessment. However, it is debated whether echocardiography should be part of the diagnostic workup in all hypertensive patients, and its routine use in all hypertensives is not endorsed by all hypertension societies.^16^ It would be ideal to expand the diagnostic capabilities of ECG, in order to detect hypertensive patients with left ventricular remodeling and LVH, and refer them for further echocardiographic evaluation.

This study was designed to test the hypothesis that a 12 lead ECG, a routine and inexpensive screening procedure, through ML methods, can provide further accuracy in detecting abnormal LVG, even at the early stages before the onset of LVH, in a population without established CVD. In addition we seek to understand which features contribute to the ML model’s decisions by calculating feature importance and feature interactions.

## METHODS

### Study population

We enrolled 528 consecutive subjects > 30 years of age with and without essential hypertension and no indications of CVD in a prospective single center study. The diagnosis of hypertension was based on the recommendations of the European Society of Hypertension/European Society of Cardiology.^15^ A physical examination and routine laboratory tests were performed before inclusion.

Patients with any of the following characteristics were excluded: pregnant or lactating women; secondary hypertension; tachy- or bradyarrhythmia; permanent atrial fibrillation, RBBB, LBBB or other conduction abnormalities on ECG, coronary artery disease; moderate or severe valvular heart disease, cardiomyopathy, cerebrovascular, liver or renal disease; history of acute coronary syndrome or myocarditis; ejection fraction < 55%; history of drug or alcohol abuse; any chronic inflammatory or other infectious disease during the last 6 months; thyroid gland disease. Vascular or neoplastic conditions were ruled out in all participants by a careful examination of the history and routine laboratory tests.

Functional tests for myocardial ischemia, coronary computed tomography angiography or invasive coronary angiography were performed according to physician’s judgement, in order to exclude coronary artery disease. A full echocardiographic examination was carried out in all participants. The study was conducted in accordance with the Declaration of Helsinki, the protocol was approved by the Hospital Ethics Committee, and patients gave written informed consent to their participation in the study.

### Echocardiography

A Standard M-mode, two-dimensional echocardiography, and Doppler measurements of LV function were performed in all subjects using a Vivid 7 (General Electric, Horten, Norway) ultrasound device with a 1.5 - 3.6 MHz wide angle phased-array transducer (M4S) according to the recommendations of the European Association of Cardiovascular Imaging and American Society of Echocardiography^17^ by two experienced echocardiographists blindly. Left ventricular internal dimension and septal and posterior wall thicknesses were measured at end-diastole, according to the American Society of Echocardiography guidelines.^16^ Abnormal LVG was identified as increased relative wall thickness (RWT) and/or left ventricular mass index (LVMi). Left ventricular hypertrophy was considered present if the left ventricular mass index was ≥ 115 *g*/*m*^2^ and 95 *g*/*m*^2^ for males and females respectively. Relative wall thickness was derived by 2 × PWT/LVIDd (LVIDd-left ventricular internal diastolic dimension, PWT-posterior wall thickness). Normal relative wall thickness was defined as < 0.43.^15^

Subjects were classified into 3 groups based on LVMi and RWT as follows: a) NG, normal geometry, defined as normal LVMi and normal RWT, b) CR, concentric remodeling, defined as normal LVMi and increased RWT, and c) LVH, concentric hypertrophy or eccentric hypertrophy, defined as increased LVMi.

### Electrocardiography

A 12-lead ECG in resting position with 10 s duration was performed in each subject using a digital 6-Channel machine (Biocare iE 6, Shenzhen, P.R. China) and were stored in XML format. To prevent ECG electrode misplacement, all ECG recordings were performed by a nurse under a cardiologist’ s supervision. Sampling rate was 1000Hz. Automated measurements of certain ECG amplitudes and durations, calculated by Biocare’s software on a 1s representative beat, were extracted from the digital files and verified by a clinician. Additional waveform measurements were extracted from the XML documents by our own custom-made software written in Python,^18^ thus producing new features.

### Clinical and anthropometric measurements

Weight and height were measured during the visit; using the World Health Organization (WHO) classification of body mass index (BMI), the individuals were classified into the three groups: normal weight (18.5 - 24.9 kg/m^2^), overweight (25 - 29.9 kg/m^2^), and obese (≥30 kg/m^2^). Body surface area (BSA) was calculated using Mosteller’s equation. The absence or presence of hypertension was documented using a binary variable (0 or 1 respectively).

### ML modeling for classification

Reservoir and ensemble machine learning methods have been applied successfully in a variety of complex physical systems^19,20^. A Random Forest (RF)^21^ is an ensemble-based supervised machine learning algorithm comprised of a collection of de-correlated decision trees. ^22^ Each decision tree performs a series of binary decisions (splits) by selecting a subgroup of input features (such as age, QTc duration, BMI class), effectively trying out different feature order and feature combinations. Each tree will give an estimate of the probability of the class label, the probabilities will be aggregated from all the trees in the RF and the highest one yields the predicted class label. RFs are good predictors even with smaller datasets due to a technique called bootstrap aggregating (bagging). Bagging trains multiple trees on overlapping, randomly selected subset of the data.

For modeling the RF we used *scikit-learn*,^23^ an open-source, powerful Python package for ML. We optimized the model parameters by minimizing the RF’s built-in *out-of-bag error* estimate which is almost identical to that obtained by N-fold cross-validation.^24^ This technique enables RFs to be trained and cross-validated in one pass.

### Predictor (feature) engineering

Our Python code extracted additional ECG waveform measurements from the 1s representative beat produced by the electrocardiograph. With the aid of automated measurements provided by the machine, we calculated several other predictors (features) such as areas under curves, slopes and peak heights of curves (Table 1). Some of the custom-engineered features are:

**Table 1.** Clinical, anthropometric, and ECG data (features) used as inputs to the machine learning (ML) model.

| Feature | Description, *Units/Classes* | Range | Mean/Mode |
| --- | --- | --- | --- |
| Sex | Male/Female | 2 | F |
| Age | Age, *years* | 31 - 90 | 61.5 |
| Hypertension | History of hypertension * | 2 | 1 |
| BMI group | Body mass index class ** | 3 | 3 |
| Height | Height, *cm* | 137.0 - 194.0 | 165 |
| BSA | Body surface area, $m^2$ | 1.3 - 2.8 | 1.93 |
| P duration | P wave duration, *ms* | 0.0 - 154.0 | 113 |
| QRS duration | QRS interval duration, *ms* | 68.0 - 138.0 | 92.7 |
| QTc duration | QT-interval corrected for heart rate ***, *ms* | 368.0 - 510.0 | 422 |
| ST duration | ST segment duration, *ms* | 0.0 - 277.0 | 112 |
| S amplitude (V1) | S wave amplitude in V1, *mV* | 0.0 - 1.9 | 0.732 |
| SL | Sokolow-Lyon voltage, *mV* | 0.5 - 4.5 | 1.93 |
| R amplitude max | Tallest R wave in limb leads, *mV* | 0.3 - 1.9 | 0.903 |
| BMI/SL | BMI divided by SL voltage, $\frac{kg}{m^2 mV}$ | 6.0 - 73.8 | 17.4 |
| ID (V5) | Intrinsicoid deflection in V5, *ms* | 24.0 - 57.0 | 39.5 |
| Area R (aVF) | Area under R wave in aVF, $ms \cdot mV$ | 0.0 - 21.3 | 3.46 |
| Area S (V1) | Area under S wave in V1, $ms \cdot mV$ | 0.0 - 62.2 | 17.3 |
| Area QRS (V5) | Area under QRS interval in V5, $ms \cdot mV$ | 11.0 - 80.2 | 33.7 |
| Area QRS (12) | Total area in all leads (sum), $ms \cdot mV$ | 137.5 - 672.9 | 285 |
| R amplitude (V2) | R wave amplitude in V2, *mV* | 0.0 - 2.0 | 0.41 |
| Cornell product | $[R(aVL)+S(V3)] \cdot \text{QRS duration, } ms \cdot mV$ | 3.6 - 243.8 | 52.6 |
| S amplitude (V2) | S wave amplitude in V2, *mV* | 0.1 - 2.3 | 0.784 |
| S amplitude (V5) | S wave amplitude in V5, *mV* | 0.0 - 1.3 | 0.381 |
| QRS-T angle | Planar Frontal QRS-T angle, *degrees* | 0.0 - 176.0 | 33.3 |
| QRS axis front | QRS axis front, *degrees* | -77.0 - 97.0 | 15.6 |
| T axis front | T axis front, *degrees* | -59.0 - 258.0 | 39.8 |
| T amplitude (aVL) | T wave amplitude in aVL, *mV* | -0.3 - 0.3 | 0.0815 |
| T amplitude (V2) | T wave amplitude in V2, *mV* | -0.3 - 1.2 | 0.253 |
| J point (V5) | J point deflection, *mV* | -0.2 - 0.1 | -0.0422 |
| R-S vector ratio | R-S vector ratio V5/V2 | 0.0 - 118.0 | 5.41 |
| S amplitude (V3) | S wave amplitude in V3, *mV* | 0.1 - 2.7 | 0.828 |
| Cornell sum | $R(aVL) + S(V3), mV$ | 0.2 - 3.8 | 1.39 |

\* 0 = no history of hypertension, 1 = history of hypertension
\*\* classes are: normal weight, overweight, obese
\*\*\* Calculated using the Bazett formula: $QTc = QT / \sqrt{RR}$ , where QTc (ms), QT (ms), $\sqrt{RR}$ (unitless)

1. Planar Frontal QRS-T angle (QRS-T angle): The difference between the axis of ventricular depolarization and repolarization,^24^ it is the absolute value of the difference between QRS axis and T-wave axis on 12-lead ECG;
2. BMI divided by the Sokolow-Lyon (SL) voltage (BMI/SL): An interaction feature, meaning a feature constructed by combining two others, BMI in continuous value divided by the sum of the amplitudes of S wave on V1 and R wave on V5.
3. Intrinsicoid deflection or R-wave peak time (ID V5): Represents the early phase of ventricular depolarization and is defined as the time period from the onset of the QRS complex to the peak of the R wave.
4. Area under the R wave measured from onset of the R wave to R peak calculated for lead aVF (R area aVF).
5. R-S vector ratio for lead V5 and V2 (R-S vector ratio): We define the vector as the sum of the R wave amplitude plus the S wave amplitude; they are opposite so this is effectively a subtraction. The R-S vector ratio is the quotient of the R-S vector in V5 divided by the R-S vector in V2.
6. QTc duration (QTc dur): QT-interval corrected for heart rate.
7. Largest R wave amplitude of the limb leads (R max).

### Feature selection

RFs are capable of handling non-linear interactions as well as correlations among features. Initially we had 60 features in our dataset. We decided to do feature selection for two reasons: the model’s accuracy improved by 4% when trained with a reduced set of features, since less irrelevant measurements were included, b) some of the features exhibited high correlation among them, as assessed by Pearson’s correlation test, so keeping only one of them retained all the information while providing a clearer picture of the remaining feature’s contribution; a Pearson’s coefficient >0.90 was our threshold for removal. Our final model was trained on 32 features (Table 1).

### Datasets

The original dataset was split into a *train set* (80%), used directly to learn the parameters of the model, and a *test set* (20%), consisting of data the model did not see during training, and was used exclusively for final performance evaluation of the models. Stratification for sex, NG class, CR+LVH class, and BMI class, done while splitting, provided *train* and *test sets* with the same proportions of these features as the original dataset. For parameter tuning during training we used RF’s internal *out-of-bag (oob)* validation. All reported performance results are on the *test set*. Feature importance graphs are also on the *test set*, as, using the *train set* inflates the importance of some features which might not be as important in predicting the outcome.

### Feature Importance

Explaining predictions from tree models is always desired and is particularly important in medical applications, where the patterns uncovered by a model are often more important than the model’s prediction performance.^25^ Scikit-learn’s tree ensemble implementation allows for the computing of measures of feature importance. These measures aspire to provide insight into which features drive the model’s prediction. Mean Decrease in Impurity (MDI), an approach popular among medical researchers, calculates each feature importance as the sum over the number of splits (across all trees). It was shown that the impurity-based feature importance can inflate the role of numerical features and bias the contribution of categorical, low cardinality ones.^26^ Furthermore, these importances are computed on training set statistics and therefore do not reflect the usefulness of the feature in predictions that generalize to the test set. A better method is Permutation Importance which randomly shuffles a feature and calculates the error after running the model; if the error increased, then that feature is deemed important. We go one step further and calculate a recent feature importance metric called SHAP (SHapley Additive exPlanations)^25,27^, a game theoretic approach to explain the output of any machine learning model. SHAP connects optimal credit allocation with local explanations using the classic Shapley values from game theory and their related extensions. Visualizing feature importance using SHAP values is thought to be more accurate for global and local feature importance (importance calculated on *each* feature instead of all of them). SHAP values have already been used in medical papers.^28^

## RESULTS

After careful screening of 903 hypertensive and normotensive healthy individuals, we enrolled 528 consecutive subjects > 30 years of age with and without essential hypertension and no indications of CVD. Of the chosen subjects, 56.0% were female, 44% were male, and 71.0% were hypertensive. The mean age was 62.3±11.9 years for women, and 60.5±12.4 years for men. Based on BMI, 45.7% of them were obese, 40.2% were overweight, and 13.9% were within normal range; there were no underweight individuals. CR was present in 37.2% of the individuals, LVH was present in 17.0%, while 45.7% of them had normal geometry. Figure 1 shows the box plots for four features and their distributions among the three categories, NG, CR, and LVH. Figure 1D indicates a tendency of BMI/SL to discriminate the NG class from the other two.

**Figure 1:**
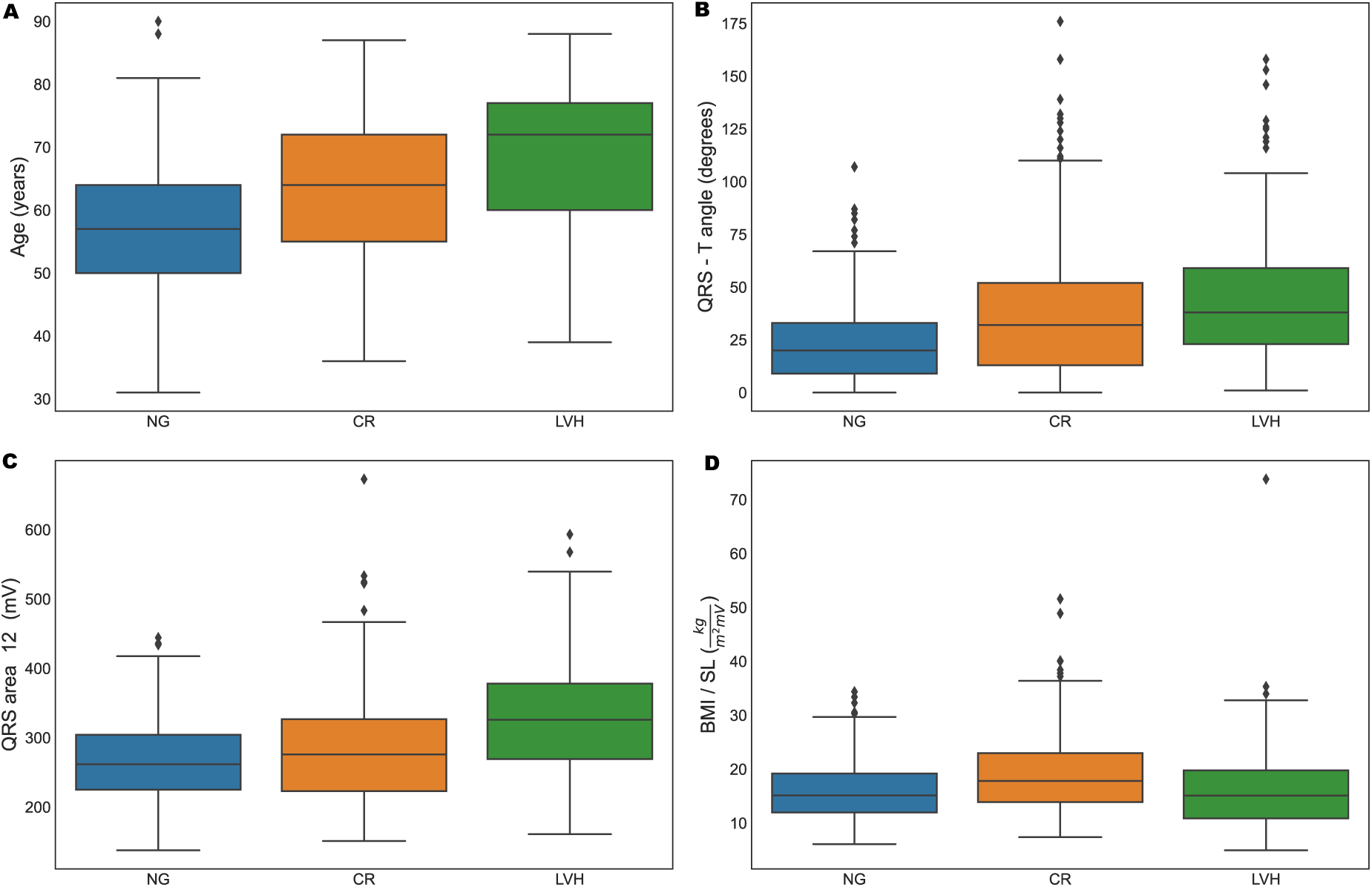
**(A) - (D)** Box plots of feature distributions in subjects for class NG, CR, and LVH separately. In figure **(D)** we notice that BMI/SL shows tendency to discriminate the CR class. The individual dots in the box plots depict outliers, patients with predictor values very different from other subjects. (BMI: body mass index, CR: concentric remodeling, LVH: left ventricular hypertrophy, NG: normal geometry)

### ML model

Trained on a set of features, an ML classifier’s goal is to assign each individual (observation) to one of various classes (response variable). We try combinations of response variables and used accuracy, sensitivity, specificity, area under the receiver operating characteristic curve (AUC/ROC), and area under the precision-recall curve (AUC/PR), for evaluating the performance of our model. The combinations are: 1. Classifying an individual as NG vs. CR+LVH (binary classification), 2. Isolating the individuals that have already developed LVH, we classify in NG+CR vs. LVH (binary classification), 3. Last, we classify to either of three classes: NG vs. CR vs. LVH (multiple classification) (Table 2).

**Table 2.** Performance metrics for the RF classifier in various categories. Imbalance correction was applied to one of them using Random Over Sampler.

| Features | Categories | Threshold | Accuracy | Specificity | Sensitivity | AUC/ROC | AUC/PR |
| --- | --- | --- | --- | --- | --- | --- | --- |
| 6 clinical | NG vs. CR+LVH | 0.50 | 0.79 | 0.73 | 0.84 | 0.85 | 0.82 |
| 6 clinical + 26 ECG | NG vs. CR+LVH | 0.50 | 0.86 | 0.75 | 0.95 | 0.89 | 0.88 |
| 6 clinical + 26 ECG +oversampling | NG+CR vs. LVH | 0.50 | 0.89 | 0.97 | 0.50 | 0.85 | 0.67 |

(AUC/PR: area under the precision-recall curve, AUC/ROC: area under the receiver operating curve, CR: concentric remodeling, ECG: electrocardiogram, NG: normal geometry, LVH: left ventricular hypertrophy)

### Binary classification NG vs. CR+LVH

Initially we trained our model using only 6 clinical variables (sex, age, BMI class, BSA, hypertension, and height), and got an accuracy of 79% in the test set, with a sensitivity of 84% and a specificity of 73%. The addition of the 26 chosen ECG-derived features improved our accuracy to 86% (in the same test set), with a sensitivity of 95% and a specificity of 75% for the default threshold of 0.5. AUC/ROC (Figure 2) was 0.89, while AUC/PR was 0.88.

**Figure 2:**
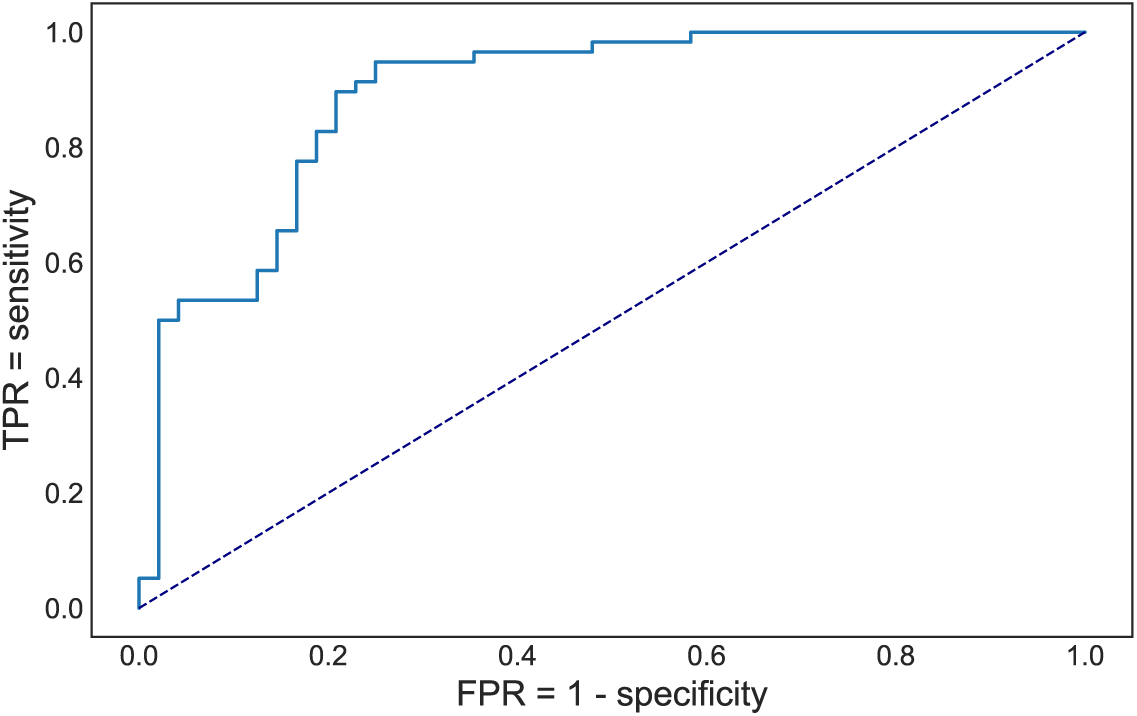
Receiver operating characteristics curve for detecting CR+LVH. True positive rate (TPR) or sensitivity, False positive rate (FPR) or (1 - specificity).

We then visualized the global feature importance and local explanations for the binary classification using SHAP. An interesting finding is the effect of specific features on each individual subject separately, as well as the interaction effects between pairs of features (Figure 3). From Figure 3B we note hypertension has a strong positive effect on being classified as CR+LVH while non-hypertensive people have different risk for being classified as CR+LVH. Age plays an important role in the risk of being classified as CR+LVH with a cut-off around 65 years of age and the risk is higher for men under 65, while over 65 the risk appears higher for women (Figure 3C).

**Figure 3:**
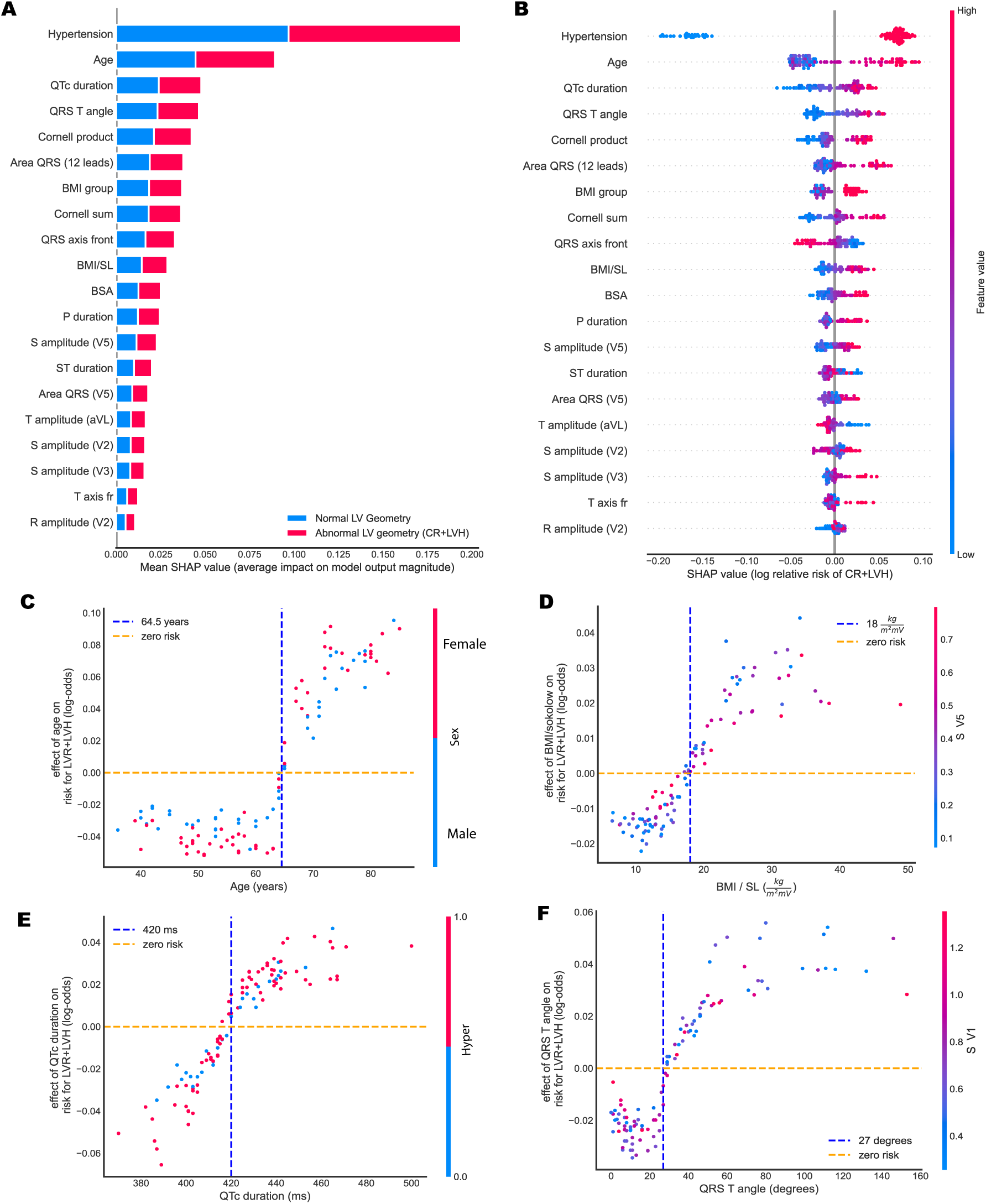
Global and local importance for the 20 most important features in the RF binary classifier for detecting CR+LVH, and feature interactions for four of them. All plots are on the test set. **(A)**: Bar chart of mean feature importance for the classification. **(B)**: SHAP summary plot showing the effect of each feature on individual patients. **(C)**: Effect of Age on detecting CR+LVH. **(D)**: Effect of the BMI/SL on detecting CR+LVH with a visible cut-off of around 18, 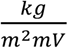. **(E)**: Effect of QRS-T angle on the same risk. Risk appears higher after a value of 27 degrees **(F)**: Effect of QTc duration on the same risk, with a visible cut-off point at 420 ms. (BMI: body mass index, BSA: body surface area, CR: concentric remodeling, ECG: electrocardiogram, LVH: left ventricular hypertrophy, NG: normal geometry, RF: random forest, SHAP: SHapley Additive exPlanations)

### Binary classification for classes NG+CR vs. LVH

Concentrating on the LVH class, we trained the RF to classify NG+CR vs. LVH. We achieved an accuracy of 89%, specificity 97%, sensitivity 50%, AUC/ROC 0.85, and AUC/PR 0.67. Due to high imbalance in the data set (ratio of NG+CR to LVH was 5/1) when divided into these categories, we performed *oversampling* for imbalance correction using *Random Over Sampler* ^*28*^ from the *scikit-learn* package.

### Multiclassification for classes NG vs. CR vs. LVH

When trained to classify subjects into three classes, NG, CR, and LVH, our model achieved an accuracy was 74%, precision 65%, and sensitivity 88% for the CR category. Distinguishing the 3 categories, enables us gain insight on the features that contribute to an individual being classified in the CR category (Figure 4). From Fig. 4D we observe that individuals with hypertension combined with high QTc duration seem to have an increased risk.

**Figure 4:**
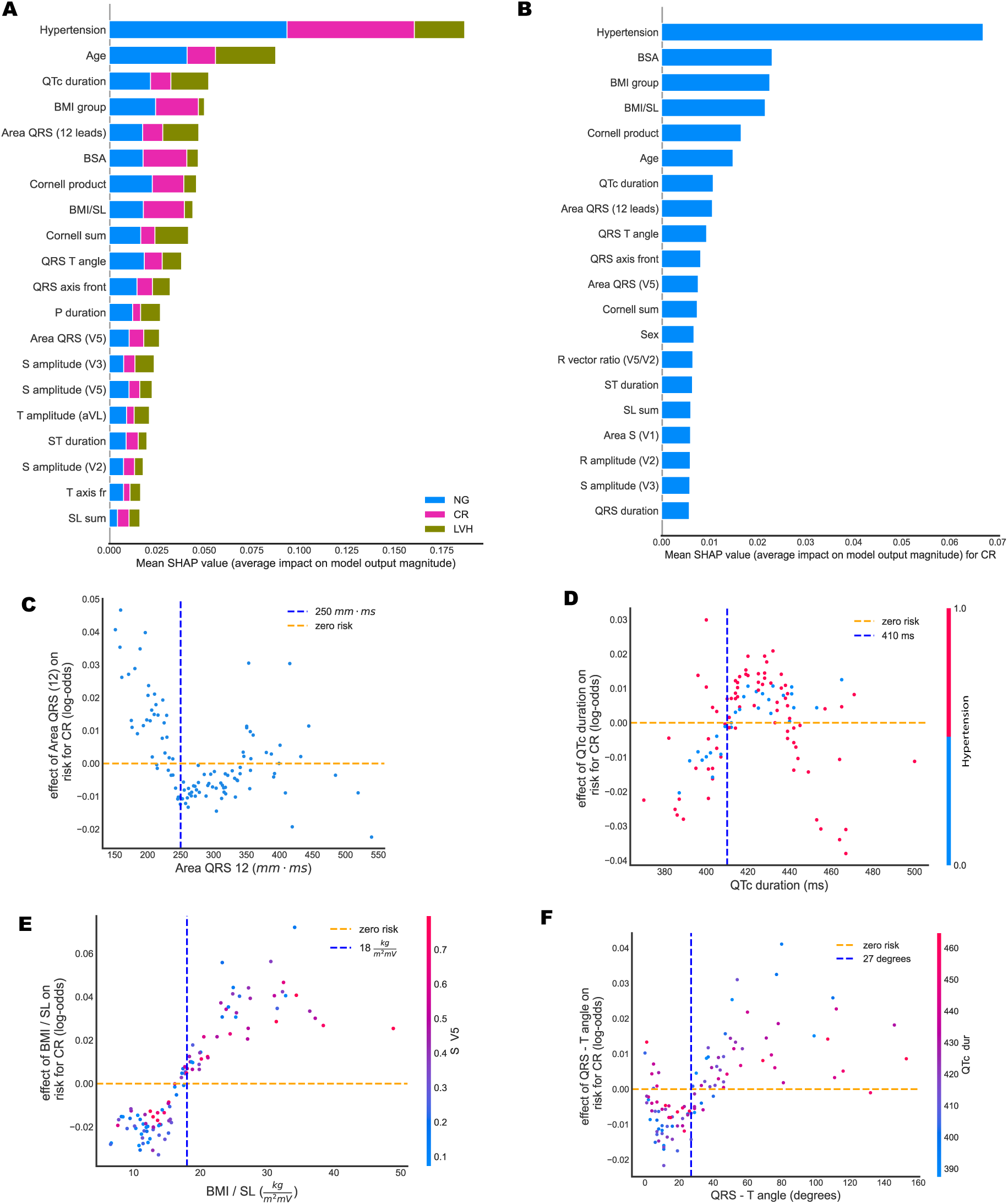
Global and local importance for 20 most important features in the RF 3-class multiclassifier, detecting NG vs. CR vs. LVH, and feature interactions for four of them. All plots are on the test set. **(A)**: Bar chart of mean global feature importance for distinguishing among the 3 classes. Colors indicate the importance of each feature to each category, with NG depicted by blue, CR by magenta, and LVH by green. **(B)**: SHAP summary plot showing the effect of each feature on detecting specifically CR. **(C)**: Effect of the area under the QRS interval summed over all 12 leads. **(D)**: Effect of QTc duration on the same risk, with a visible cut-off point of 410 ms. **(E)**: Effect of the BMI/SL on detecting CR with a visible cut-off around 17 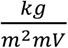. **(F)**: Effect of QRS-T angle on the risk of having CR. (BMI: body mass index, BSA: body surface area, CR: concentric remodeling, ECG: electrocardiogram, LVH: left ventricular hypertrophy, NG: normal geometry, RF: random forest, SHAP: SHapley Additive exPlanations)

## DISCUSSION

To our knowledge, our study is the first to demonstrate the promising potential of ML modeling for the efficient and cost-effective diagnostic screening of abnormal LVG and cardiac remodeling through ECG. We found specific clinical and ECG features that can predict early pathological changes of LVG in patients without established CVD and detect the population who will benefit from a detailed echocardiographic evaluation. We used not only the traditional ECG criteria for LVH but also novel ECG markers that increased the accuracy of our ML model. Our findings are very significant given that a patient population is included that have LV remodeling at very early stages without LVH, a situation that until now was not detectable by ECG but required imaging methods.

We know that risk factors such as arterial hypertension might influence LV morphology and increase in LV volumes and sphericity, increase LV mass, compensate LV function which finally compromises stroke volume and ejection fraction. Identification of LV geometric patterns is substantial.^14^ Longstanding or inadequately treated hypertension will result in changes in LV shape and eventually, a deterioration of systolic function.

The detection of hypertension-mediated organ damage, such as abnormal LVG, is a useful approach toward risk stratification of a hypertensive population.^15^ The evaluation of cardiac structure and function is encouraged since it might influence treatment decisions.^15^ Transthoracic echocardiography has received a strong indication for the initial evaluation of suspected hypertensive heart disease. Abnormal LVG is the early marker of LV remodeling that precedes hypertrophy and is frequently associated with LV diastolic dysfunction.^17^ In hypertensive patients, the type of LVG and LV remodeling (CR, eccentric and concentric LVH) is predictive of the incidence of cardiovascular (CV) events.^15^ CR not only precedes LVH but also most other cardiac dysfunctions while it progresses asymptomatically. The early detection of abnormal LVG can result in early detection of subclinical hypertension-mediated organ damage and may help clinical decision and follow up.

ML classifiers, specifically RFs, trained on clinical information and measurements from the ECG, one of the most common non-invasive diagnostic techniques, identify normal individuals, versus ones that have either LVH or LV remodeling at initial stages. Many studies confirm that ensemble methods outperform any single base learner such as Classification and Regression Decision Trees (CART). For cases of complex datasets, such as the one we have, linear-based algorithms such as Logistic Regression may not be sufficient in segmenting the class labels, leading to poor accuracies. More sophisticated algorithms such as random forests, which can learn a non-linear decision boundary, are more effective and can achieve higher accuracy scores.

Our findings show that age plays an important role in the risk of someone having CR or LVH with a cut-off around 64.5 years. The risk appears higher for men younger than 64.5 while after that age the risk seems higher for women. We also introduce the quotient of BMI and the BMI/SL because we hypothesize that body mass affects the amplitude of the R and S waves, as the electrical currents cover different distances. Our results indeed indicate that BMI/SL seems to differentiate for the CR class. Hypertension, age, and BMI were most significant, as expected; the area under the QRS complex summed over all 12 leads, the Planar Frontal QRS-T angle, and QTc duration, among others, were important in predicting risk.

There are limited data in the literature that attempt to predict cardiac structural or functional abnormalities with ECG data interpreted through ML algorithms.^30–32^ However, the existing knowledge has focused only on patients who have already shown LVH. There are no data for patients in earlier stages of cardiac geometry change prior to hypertrophy. The present prospective ML study also differs from previous ones in that it involves patients who were very carefully selected, thereby excluding those with CVD.^30,31^ This may explain the fact that in our study, analysis of patients with LVH achieved a higher AUC in comparison to recently published work,^30^ despite the fact that the number of our patients is smaller. ML is susceptible to major errors in interpretation, and generalizability. The fact that participants in our study did not have CVD is a major strength since in effect it largely eliminates other clinical parameters that could mislead our model. In this way, we improve the quality of input data and avoid various pitfalls that could arise due to the large diversity of pathological conditions that formed the basis for the training process.

We have discovered new relationships and showed that a quantitative assessment of abnormal LVG can be performed by using easily obtained clinical data and ECG features. This novel approach has the potential to serve as a cost-effective screening tool for early detection of LV remodeling, to dramatically optimize treatment and patient-management. ECGs are more easily obtainable and cost-effective than echocardiography or cardiac magnetic resonance imaging (MRI), and for those reasons more often used in current clinical practice. The goal of hypertension treatment is to prevent pathological changes in LVG or to reverse CR and LVH. Deep learning could potentially detect patients with hypertension-mediated organ damage at an early stage and with simple and widely used clinical tools. Digital health care opens up new opportunities in health care quality and advancing personalized medicine at a lower cost. We showed that from basic clinical data and the use of ECG, we can distinguish high risk patients such as the ones beginning to show CR; these are the ones requiring further evaluation, closer follow up and more detailed cardiovascular imaging. This initial approach can be performed in primary care facilities, or even out of office. Our model contributes to the development of human-centered and autonomous technologies and can optimize patient-management and treatment. This has become crucially important in recent times, due to the unprecedented demands on healthcare systems worldwide that the pandemic outbreak has imposed.

### Limitations

The number of subjects we have included is not large. Nonetheless, our results are clear and mainly due to the fact that our subject population is carefully chosen and does not have other CVD that could influence ECG features.

Although cardiac MRI has been suggested as the most accurate diagnostic test for cardiac remodeling and LVH, it is limited by cost and lack of availability. Most importantly, it is not recommended for routine clinical use for this reason. 2D echocardiography is the imaging test of choice for assessing those patients and is also the only guideline-approved modality for monitoring volumes and mass,^15,17^ which also has well studied prognostic and clinical implications.^12,15^

We did not perform coronary angiogram in all patients, and this may bias the outcomes. However, we believe that this bias is small since patients underwent a meticulous work out to exclude coronary artery disease while performing coronary angiogram in low probability patients would be unethical.

RWT is not always reflective of true LVG in patients with asymmetric hypertrophy. However, it is the most widely used index for this purpose in routine clinical practice for hypertensive patients.^15,17^ More studies are needed to test the applicability and transferability of our patients to other patients’ cohorts.

### Perspectives

We developed novel ML algorithms that are effective in the detection of patients with abnormal LVG even at very early stages, before the progression to LVH. Hypertension, age, BMI over the Sokolow-Lyon voltage, QRS-T angle, and QTc duration were some of the most important features for this purpose. Our method offers an innovative strategy to improve health care management and personalized care at lower cost, especially in patients at risk for CVD, such as the hypertensive population. Further studies are required to determine whether our criteria can be broadly applied to other populations. Although there are still challenges in ML-based applications to cardiology, research should be expanded since ML-models can efficiently identify actionable insights into disease processes.

## Data Availability

The data underlying this article cannot be shared publicly due to the privacy of individuals that participated in the study. The data will be shared on reasonable request to the corresponding author.

## FUNDING

This work was partial supported by the Institute of Theoretical and Computational Physics of the University of Crete.

## ACKNOWLEDGEMENTS

We thank the Center for Quantum Complexity and Nanotechnology of the University of Crete for providing access to the Metropolis Supercomputer. One of us (E. Angelaki) would like to thank Dr. Pavlos Protopapas (IACS, Harvard University) for his feedback during the development of the ML models.

## CONFLICT OF INTEREST

Professor Vardas has to report personal fees from MENARINI INTERNATIONAL, DEAN MEDICUS, SERVIER, EUROPEAN SOCIETY OF CARDIOLOGY, HYGEIA HOSPITALS GROUPS LTD, BAEYR. The other authors have nothing to declare.

### Summary

We used ML techniques and found a combination of clinical and ECG features that enable prediction of abnormal LVG or LVH. CR appears to be separable from LVH through a RF method.

## Notes

### Author Declarations

The study was carried out in accordance with the Declaration of Helsinki, the protocol was approved by the Hospital Ethics Committee and the Research Ethics Committe of the University of Crete. (Ν.4521/2018) All patients gave written informed consent to their participation in the study.

### Summary of Updates

One sentence addition to Acknowledgements

